# Year-long COVID-19 infection reveals within-host evolution of SARS-CoV-2 in a patient with B cell depletion

**DOI:** 10.1101/2021.10.02.21264267

**Authors:** Veronique Nussenblatt, Allison E Roder, Sanchita Das, Emmie de Wit, Jung-Ho Youn, Stephanie Banakis, Alexandra Mushegian, Christopher Mederos, Wei Wang, Matthew Chung, Lizzette Pérez-Pérez, Tara Palmore, Jennifer N. Brudno, James N. Kochenderfer, Elodie Ghedin

**Author notes:** Contributed equally to this article (Veronique Nussenblatt, Allison E Roder).

## Abstract

**Background:** B-cell depleting therapies may lead to protracted disease and prolonged viral shedding in individuals infected with SARS-CoV-2. Viral persistence in the setting of immunosuppression raises concern for viral evolution.

**Methods:** Amplification of sub-genomic transcripts for the E gene (sgE) was done on nasopharyngeal samples over the course of 355 days in a patient infected with SARS-CoV-2 who had previously undergone CAR T cell therapy and had persistently positive SARS-CoV-2 nasopharyngeal swabs. Whole genome sequencing was performed on samples from the patient’s original presentation and 10 months later.

**Results:** Over the course of almost a year, the virus accumulated a unique in-frame deletion in the amino-terminal domain of the spike protein, and complete deletion of ORF7b and ORF8, the first report of its kind in an immunocompromised patient. Also, minority variants that were identified in the early samples—reflecting the heterogeneity of the initial infection—were found to be fixed late in the infection. Remdesivir and high-titer convalescent plasma treatment were given, and the infection was eventually cleared after 335 days of infection.

**Conclusions:** The unique viral mutations found in this study highlight the importance of analyzing viral evolution in protracted SARS-CoV-2 infection, especially in immunosuppressed hosts, and the implication of these mutations in the emergence of viral variants.

**Summary:** We report an immunocompromised patient with persistent symptomatic SARS-CoV-2 infection for 335 days. During this time, the virus accumulated a unique in-frame deletion in the spike, and a complete deletion of ORF7b and ORF8 which is the first report of its kind in an immunocompromised patient.

## BACKGROUND

Cell-mediated and humoral immunity are necessary to clear SARS-CoV-2 infection [1, 2]. Individuals receiving B-cell depleting therapies can have protracted disease and prolonged viral shedding [3-6]. Persistent shedding of viral RNA for weeks to months after onset of symptoms has been reported, however viable virus is not detected after 9 days post illness onset in most patients [7]. In contrast, viral replication has been detected in immunocompromised patients for several months after initial infection [5, 8, 9]. Persistent viral replication in these patients is likely the result of profound lymphocyte defects due to B- and T-cell depleting therapies or underlying hematologic disease. Viral persistence in the setting of immunosuppression has raised concern for viral evolution [9] and the emergence of variants, especially during treatment with convalescent plasma [6].

Recent studies have demonstrated that SARS-CoV-2 in immunocompromised hosts is prone to significant deletion mutations in the spike protein, especially in the S1 region [5, 8, 9]. Deletions across the genome can reflect virus-host interactions and are found in both immunocompetent and immunosuppressed hosts. For example, accessory proteins ORF7ab and ORF8, which are associated with early stimulation of IFN-γ production [10], were found to be deleted in a substantial number of immune competent individuals.

Here, we report a patient with persistent symptomatic viral infection over a period of at least 335 days. Viral genome sequencing revealed the emergence of a unique in-frame deletion in the amino-terminal domain (NTD) of the spike protein, and a complete deletion of the ORF7b and ORF8 coding regions; such a large deletion of the ORF7b-ORF8 region of the genome is the first report of its kind in an immunocompromised patient. Remdesivir and high-titer convalescent plasma treatment were administered after the appearance of deletion mutations in the virus genome, with subsequent clearance of the infection.

## METHODS

### Approvals

Written consent was obtained for human research subjects, as approved by the NIH Institutional Review Board (protocol # NCT02659943). The patient consented to have the results of this research published.

### SARS-CoV-2 RNA and sgRNA qPCR

Detection of the N gene or ORF1a/b was performed as part of diagnostic testing on all specimens collected. Amplification of sub-genomic transcripts for the E gene (sgE) was done prospectively on samples after day 275 and retrospectively on 19 samples prior to day 275, following methods described previously [8].

### SARS-CoV-2 amplification, library preparation and sequencing

Amplification of viral genomes was done with custom tiling primers using a modified version of the ARTIC consortium protocol for nCoV-2019 sequencing (https://artic.network/ncov-2019) and the methods described in *Roder et al 2021* [15]. All libraries were prepared using the Nextera XT library preparation kit (Nextera), scaled down to 0.25x of the manufacturer’s instructions and libraries were sequenced on the Illumina NextSeq500 using the 2×300 bp paired end protocol.

### Genome assembly, generation of consensus sequences and identification of variants

Following sequencing, Illumina sequencing adapters and primer sequences were trimmed with Trimmomatic v0.36 [16]. The trimmed reads were aligned to the Wuhan-Hu-1 SARS-CoV-2 reference genome (NC_045512.2) using BWA mem v0.7.17 with the -K parameter set to 100000000 for reproducibility and -Y to use soft clipping for supplementary alignments [17]. The two primer pool libraries for each biological sample were merged into one alignment file using Picard Tools MergeSamFiles v2.17.11. Duplicates were marked using GATK MarkDuplicatesSpark v4.1.3.0 (https://gatk.broadinstitute.org/hc/en-us/articles/360037224932-MarkDuplicatesSpark). The pipeline used to analyze the data is available at https://github.com/gencorefacility/covid19. Consensus sequences, consensus mutations and minority variants were called using an in-house variant calling pipeline, *timo*, which is available at https://github.com/GhedinLab/timo. Minority variants were called if present at or above an allele frequency of 0.02 and a coverage of 200X.

### Generation of phylogenetic trees and lineage identification

Phylogenetic trees were generated using the Nextstrain software package and default parameters [18]. A total of 266 sequences from Maryland collected between April 2020 and March 2021 were downloaded from GISAID and included as background and are available in supplementary table 1 [19]. Lineages were called using Nextclade and Pangolin software packages [18, 20, 21].

## RESULTS

### Case presentation

A woman in her 40’s with type II diabetes mellitus and diffuse large B-cell lymphoma (DLBCL), who had been in complete remission for three years, presented with fever, headache, nasal congestion, and productive cough. She reported that her symptoms developed eleven days prior to presentation. The patient’s history is relevant for treatment with multiple lines of therapy and anti-CD19 chimeric antigen receptor-modified T-cell (CAR-T) therapy [11] three years prior, resulting in ongoing B-cell aplasia, hypogammaglobulinemia, CD4 lymphopenia, and recurrent upper respiratory infections. A previous attempt at intravenous immunoglobulin therapy (IVIG) replacement was complicated by infusion reactions, after which the patient intermittently declined prophylactic IVIG.

A chest computerized tomography (CT) exam performed on admission showed scattered, bilateral, ground-glass radiodensities and consolidations and she required 2L of supplemental oxygen via nasal cannula (NC). Laboratory evaluation revealed a white blood cell count of 4.67 × 10^9^ cells/L (3.98 × 10^9^ cells/L-10.04 × 10^9^ cells/L), absolute lymphocyte count of 0.81 × 10^9^ cells/L (1.18 × 10^9^ cells/L-3.74 × 10^9^ cells/L), absolute neutrophil count of 3.41 × 10^9^ cells/L (1.56 × 10^9^ cells/L-6.13 × 10^9^ cells/L), IgG of 144 mg/dL (700-1600mg/dL), IgM 12 mg/dL (40-230 g/dL), IgA 31mg/dL (70-400mg/dL), CD4 count of 202/mcl. Nasopharyngeal (NP) swabs were negative for SARS-CoV-2 PCR on three occasions prior to presentation. Her symptoms worsened and she required 5L of supplemental oxygen via NC. Due to her worsening clinical status, bronchoalveolar lavage (BAL) was performed on Day 1 (May 2020). Broad microbiological testing of the BAL fluid was negative, except a positive PCR test for SARS-CoV-2. The patient’s supplemental oxygen requirement increased to 60% FiO_2_ and flow of 40 liters/minute (LPM) via high flow nasal cannula and a vasopressor was initiated, in addition to broad spectrum antibiotics. On Day 2, she received convalescent plasma and 40g of 10% immune globulin IV for her underlying hypogammaglobulinemia. She did not receive remdesivir as it was not available at the time of initial disease presentation. Corticosteroids were not administered as robust clinical trial data regarding their use in the acute setting of COVID-19 was not yet available.

The patient was discharged a month later with temperatures of 99-100°F, episodes of worsening cough and 3L NC supplemental oxygen. Testing for SARS-CoV-2 by PCR on NP swabs was performed monthly for 3 months and every 3 months, thereafter. These were positive intermittently with Ct values ranging between 37 and 38 (**Fig. 1**). Due to the patient’s overall mild to absent symptoms, positive SARS-CoV-2 tests during this period were thought to probably reflect shedding of non-viable virus particles; however, repeat chest CTs over the same period showed bilateral increasing multifocal ground-glass opacities with crazy paving pattern and mixed changes. At this time, organizing pneumonia and superimposed bacterial or fungal infection were considered. The patient preferred conservative management and declined bronchoscopy to rule out a superimposed infection. Induced sputum was negative for bacterial, fungal, or mycobacterial pathogens.

**Figure 1.**
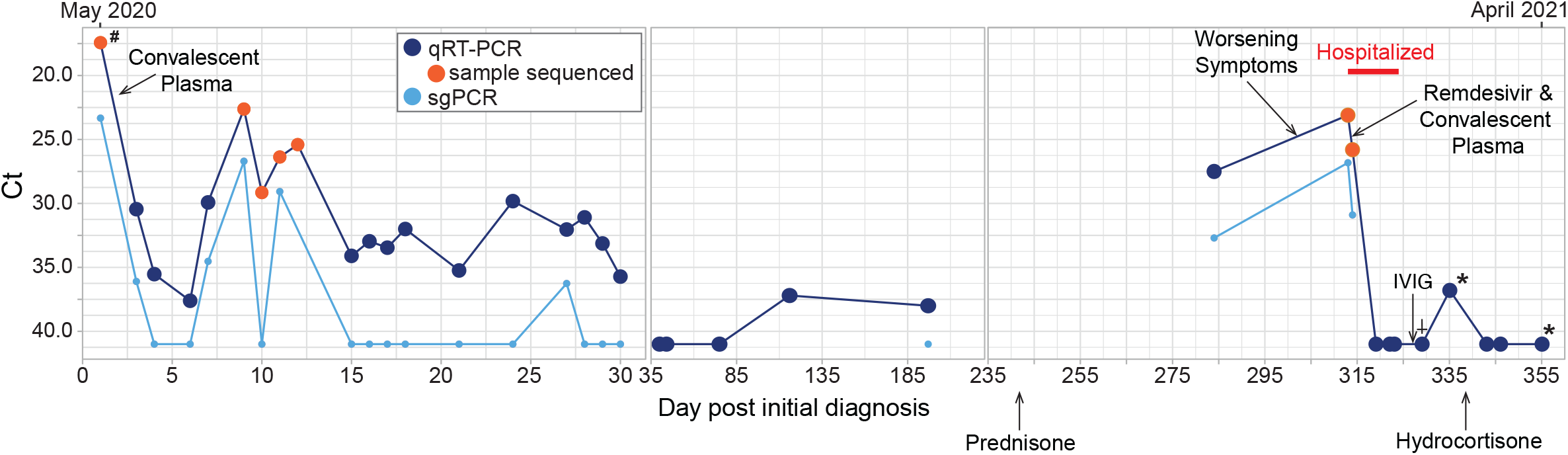
Timeline of diagnostic tests for SARS-CoV-2 and treatment. Nasopharyngeal or oropharyngeal (upper respiratory) specimens were collected for detection of SARS-CoV-2 RNA, except when indicated by the following: ^#^ indicates BAL sample, * indicates days when sputum specimens were collected and † indicates days when saliva was collected. Specimens with Ct values over 40 were considered negative for SARS-CoV-2 RNA. PCR for sub-genomic RNA was performed only on specimens that tested positive for genomic RNA. Samples that were used for next-generation sequencing are indicated with an orange circle. Treatments administered are indicated with a black arrow and labeled.

On Day 242, prednisone 50mg daily was initiated for the treatment for COVID-19-related cryptogenic organizing pneumonia and resulted in moderate symptom and slight radiographic improvement. SARS-CoV-2 PCR from a NP sample on day 284 was positive with a Ct value of 27.5, a marked decrease from the Ct value of 38 on day 197, indicating a substantial increase in viral load. This viral load increase in the setting of steroids and only modest decrease in symptoms was concerning for SARS-CoV-2 relapse versus new infection. A Ct value of 32.7 from sub-genomic RNA (sgRNA) real-time PCR performed at this time indicated recent virus replication [12-14] (**Fig. 1**). SARS-CoV-2 antibody testing was also performed but was negative. Shortly after, the patient reported worsening of respiratory symptoms with new oxygen requirement of 6L NC at rest. C-reactive protein (CRP) had been 37-80mg/L (<3.0 mg/L) and rose to 144mg/L after prednisone initiation. She was admitted to the hospital on Day 313 and treated with high-titer convalescent plasma and a 10-day course of remdesivir. She was finally discharged on Day 324 (March 2021) on 3-4L/min of oxygen at rest and 6L/min with exertion. Prednisone was tapered to physiologic doses of hydrocortisone. Three months later, CRP had normalized, CT chest showed significant decrease in ground glass opacities and the patient no longer needed supplemental oxygen at rest.

### Genomic analyses

Since diagnostic testing for SARS-CoV-2 indicated high viral load in this patient 10 months after initial diagnosis, we did whole genome sequencing on five samples from the patient’s original presentation and two samples from her second presentation to determine if re-infection had occurred. Assembled consensus sequences of virus genomes were assigned lineage and all samples from this patient mapped to the Pango lineage B.1.332 (Nextclade lineage 20C). Global surveillance of SARS-CoV-2 genomes reveals that B.1.332 was circulating at the time of her initial presentation but was no longer prevalent by the time of her second presentation [20, 21]. Consensus sequences were mapped onto a phylogenetic tree containing 266 background samples from Maryland from the time of the first to the second presentation using the publicly available Nextstrain software package [18]. All samples from this patient clustered on the same branch of the tree, with no intermixed background samples, indicative of a prolonged infection over 335 days, rather than a re-infection event (**Fig. 2**).

**Figure 2.**
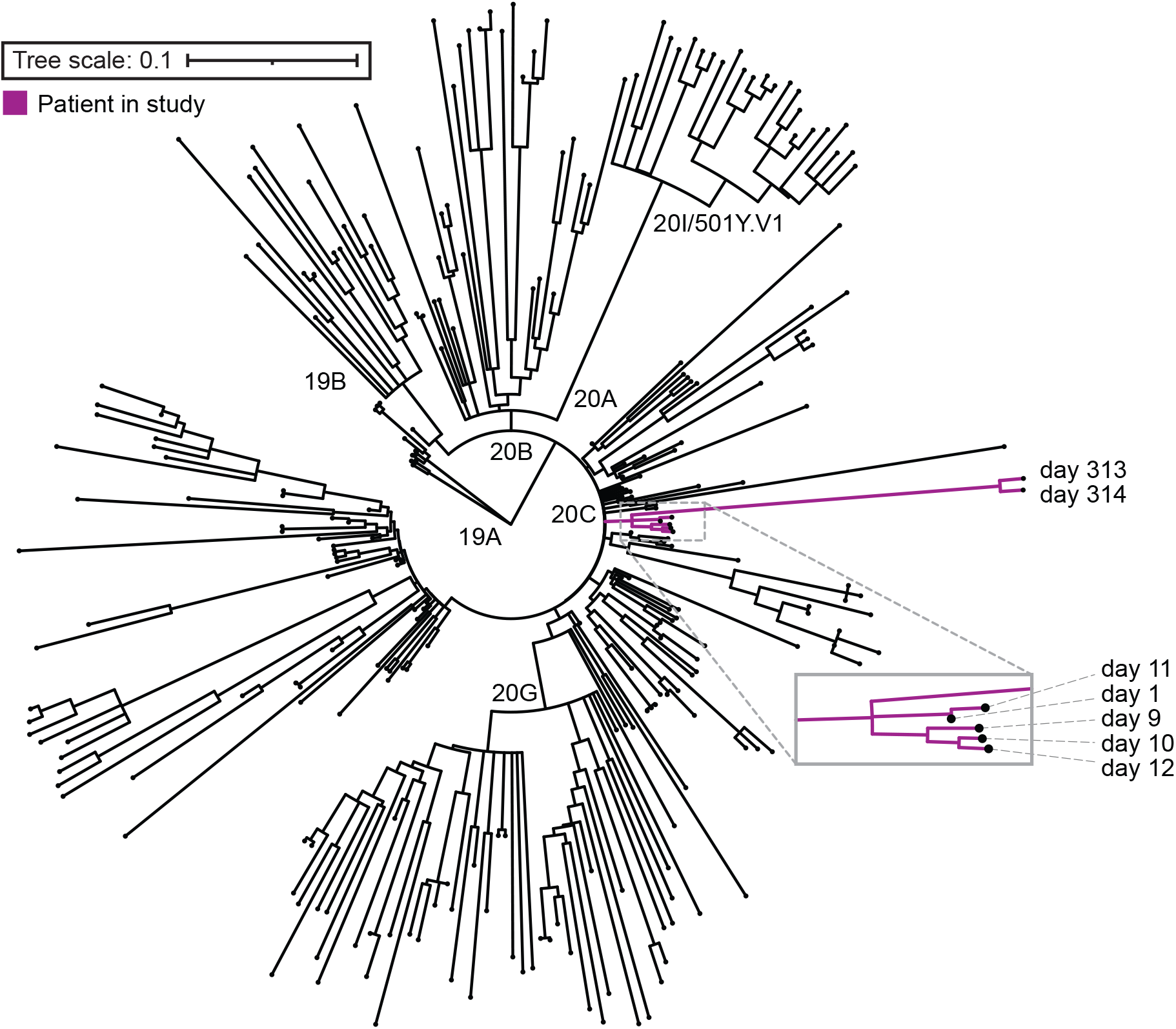
Phylogeny of sequenced samples. Maximum likelihood timed strain tree reconstructed from 266 local sequences from GISAID (**Table S1**). Samples sequenced from the patient in this case study are colored in purple and labeled with the day of infection from first diagnosis.

The original sample, taken on Day 1, contained 11 consensus changes from the Wuhan/Hu-1 strain (NC_045512.2). To visualize evolution of the virus over time within the patient, we compared the consensus nucleotides in the later 6 samples to that of the first sample (Day 1). Other samples collected the first month of infection had between 1-5 consensus changes compared to the initial sample, whereas the later samples (Days 313-331) had 28 (Day 313) and 26 (Day 314) consensus changes at the nucleotide level from the initial sample. Of those, 19 and 17, respectively, were non-synonymous, with four substitutions in the spike protein (**Fig. 3A**). More interestingly, the later samples contained 2 deletions: a gap at nt 22290 to 22298 that led to a unique del244-246 and, consequently, a A243G substitution (**Fig. 3B**); and a 497nt deletion spanning the entire length of the ORF7b coding region and all but two amino acids of the ORF8 (**Fig. 3C**). Of note, several of the amino acid changes identified in the later samples (days 313 and 314) were present as minority variants in the initial samples, suggesting a heterogeneous infection early on (**Fig. 3D**, smaller circles) with eventual fixation, as observed for ORF1a: A3070V, ORF7a: S37F, and N: P365L. Conversely, a consensus change present in the early samples also existed as minority variant in the last sample on Day 335 (**Fig. 3D**, smaller circles). The observed number of consensus changes in the later samples from the initial sample indicates that the virus evolved within this patient at approximately the same evolutionary rate that has been reported for SARS-CoV-1 [22], and for SARS-CoV-2 in the global population [23], estimated to be around 2 fixed mutations per month.

**Figure 3.**
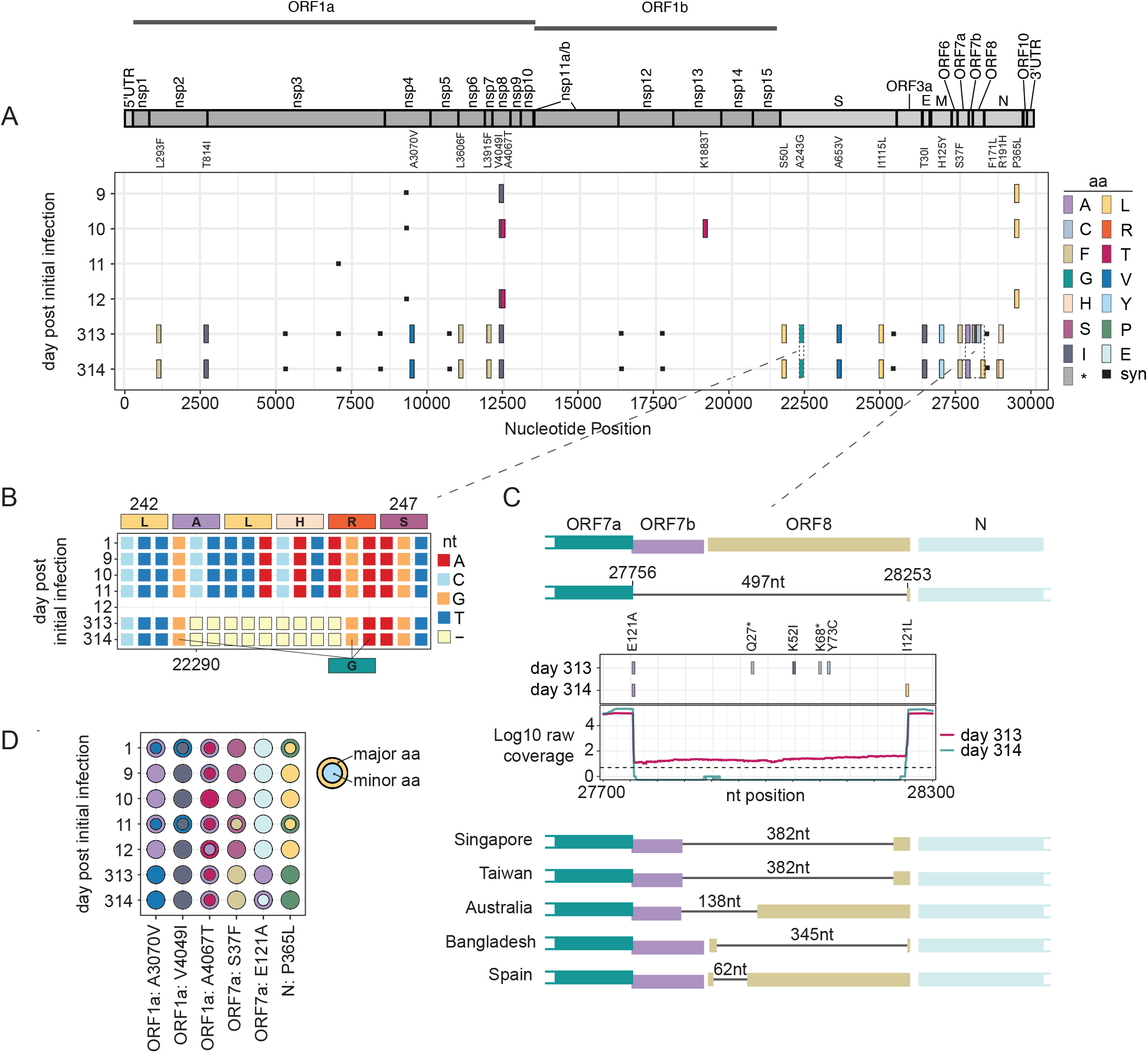
Mutations and deletions in sequenced samples over time. **(A)** Tile plot showing consensus changes across the genome as compared to the initial infectious sample (collection date: 2020-05-01, day 1), color-coded by residue. **(B)** Schematic showing 9 nt deletion in Spike region (ntpos: 22290-22299, spike aa positions shown). **(C)** Coverage plot, tile plot and schematic showing the 497nt deletion in the ORF7b and ORF8 coding regions. Coverage plot represents log10 raw coverage. Dotted line is at 5X coverage. Tile plot represents amino acid changes in this region, as compared to initial infectious sample (day 1), colored by residue as above in 3A. Deletions identified in previous studies are shown as schematics and labeled by their country of origin. **(D)** Circle plot showing major (larger, outer circle) and minor (smaller, inner circle) at locations where a minority variant in one sample exists as a major amino acid in another sample, colored by residue as in 3A. ORFs and aa positions within the encoded protein are listed.

## CONCLUSIONS

The sequencing data clearly indicate that the patient exhibited a prolonged infection over a 335-day period, the longest reported infection with SARS-CoV-2 to date. The existence of ongoing infection is further illustrated by significant clinical improvement and normalization of CRP after viral clearance, and the fact that the virus has largely followed an evolutionary trajectory that would be expected based on the mutation rate of the polymerase. While the patient displayed COVID19-like symptoms over the entirety of the infection, lower respiratory samples that may have demonstrated ongoing viral replication in between the patient’s two admissions were not available. Consequently, we cannot definitively attribute all the lung findings to infection.

Over the course of the 335-day infection, the virus accumulated mutations at approximately the same rate as expected based on the error rate of the polymerase. This was not surprising given that in both immunocompetent and immunocompromised hosts the error rate of the polymerase is likely to remain constant, though which mutations are selected for may differ due to the lack of immune pressure on the virus. Two important deletions were identified in the later samples, one in the spike protein, and one in the ORF7b and ORF8 regions. The specific NTD deletion in the spike protein that targeted residues at positions 244-246—a range that appears unique, especially for an early lineage—would impact the supersite and could induce resistance against NTD-directed antibodies [24]. This type of deletion has also been observed in variant B.1.351 (Lambda), which contains NTD deletion 242-244 and a R246I mutation [24]. It is unclear why a deletion in that region of the NTD would appear in an immunocompromised patient, but it supports previous observations where chronic SARS-CoV-2 infection in severely immunocompromised hosts receiving convalescent plasma can lead to variant emergence and reduced sensitivity to neutralizing antibodies [6]. Additionally, our patient received convalescent plasma during her first admission

The 497nt deletion in the ORF7b and ORF8 genes is the longest deletion reported in this region of the genome, and the first to be observed in an immunocompromised patient. Other reported ORF7b/ORF8 deletions in this region span from 62nt to 382nt, with the first instance identified in Singapore in January of 2020 [25, 26]. *In vitro* analyses of similar deletions indicated that deletion mutants had higher titers but similar levels of cytopathic effect and showed that the virus was still transmissible. However, the deletion mutant may be less effective at establishing infection in a new host due to loss of immune evasion features of ORF8 [26]. ORF8 has been established as a key antagonist of innate immunity and has been shown to elicit a robust and highly specific immune response during infection, suggesting that the deletion in competent hosts may be due to immune driven selection [27]. Thus, it was surprising to see emergence of this large deletion in our immunocompromised patient. It is possible that the immunocompromised nature of this patient removes a need for ORF8 during infection. This hypothesis is supported by data that show ORF8 is particularly tolerant to mutation, acquiring many missense and nonsense mutations. ORF8 was also shown to be dispensable in cell culture [28]. A retrospective cohort study performed on patients in Singapore found that some patients carried a mix of wild-type and a 382nt deletion mutant in the ORF7-ORF8 region, while others only had the deletion mutant. Over time, the deletion mutant outcompeted the non-deletion virus [29]. In our study, we also found evidence of a mixed infection with deletion and non-deletion mutants in the Day 313 sample, but infection with only the deletion mutant on Day 314, indicating that the same competition may have occurred in this patient with rapid clearance of the wild-type virus (**Fig. 3C**).

This case demonstrates that severely immunocompromised patients may experience protracted SARS-CoV-2 infection with mild symptoms and persistent virus replication. Our case also highlights the limitations of SARS-CoV-2 testing, especially for lower respiratory tract disease, as access to a BAL specimen may have alerted us earlier to ongoing infection. Further research is necessary to understand the evolution of SARS-CoV-2 in immunocompromised hosts, especially in relation to implications for viral transmission and variant emergence.

## Data Availability

Yes

## ACKNOWLEDGEMENTS

This work was funded in part by the Division of Intramural Research (DIR) at the National Institute of Allergy and Infectious Diseases of the NIH. We thank the Department of Laboratory Medicine staff for processing clinical specimens and providing technical assistance. This work utilized the computational resources of the NIH HPC Biowulf cluster (http://hpc.nih.gov).

## DATA AVAILABILITY

Data is available in NCBI GenBank under the following accession numbers: MZ385697-MZ385702 and MW990333.

## REFERENCES

1. Zohar, T. and G. Alter, *Dissecting antibody-mediated protection against SARS-CoV-2*. Nat Rev Immunol, 2020. 20(7): p. 392–394.

2. Sekine, T., et al., *Robust T Cell Immunity in Convalescent Individuals with Asymptomatic or Mild COVID-19*. Cell, 2020. 183(1): p. 158–168 e14.

3. Karatas, A., et al., *Prolonged viral shedding in a lymphoma patient with COVID-19 infection receiving convalescent plasma*. Transfus Apher Sci, 2020. 59(5): p. 102871.

4. Kenig, A., et al., *Treatment of B-cell depleted COVID-19 patients with convalescent plasma and plasma-based products*. Clin Immunol, 2021. 227: p. 108723.

5. Hensley, M.K., et al., *Intractable COVID-19 and Prolonged SARS-CoV-2 Replication in a CAR-T-cell Therapy Recipient: A Case Study*. Clin Infect Dis, 2021.

6. Kemp, S.A., et al., *SARS-CoV-2 evolution during treatment of chronic infection*. Nature, 2021. 592(7853): p. 277–282.

7. Cevik, M., et al., *SARS-CoV-2, SARS-CoV, and MERS-CoV viral load dynamics, duration of viral shedding, and infectiousness: a systematic review and meta-analysis*. Lancet Microbe, 2021. 2(1): p. e13–e22.

8. Avanzato, V.A., et al., *Case Study: Prolonged Infectious SARS-CoV-2 Shedding from an Asymptomatic Immunocompromised Individual with Cancer*. Cell, 2020. 183(7): p. 1901–1912 e9.

9. Choi, B., et al., *Persistence and Evolution of SARS-CoV-2 in an Immunocompromised Host*. N Engl J Med, 2020. 383(23): p. 2291–2293.

10. Tan, A.T., et al., *Early induction of functional SARS-CoV-2-specific T cells associates with rapid viral clearance and mild disease in COVID-19 patients*. Cell Rep, 2021. 34(6): p. 108728.

11. Brudno, J.N., et al., *Safety and feasibility of anti-CD19 CAR T cells with fully human binding domains in patients with B-cell lymphoma*. Nat Med, 2020. 26(2): p. 270–280.

12. Wolfel, R., et al., *Virological assessment of hospitalized patients with COVID-2019*. Nature, 2020. 581(7809): p. 465–469.

13. Kim, D., et al., *The Architecture of SARS-CoV-2 Transcriptome*. Cell, 2020. 181(4): p. 914–921 e10.

14. Speranza, E., et al., *Single-cell RNA sequencing reveals SARS-CoV-2 infection dynamics in lungs of African green monkeys*. Sci Transl Med, 2021. 13(578).

15. Roder, A.E., et al., *Diversity and selection of SARS-CoV-2 minority variants in the early New York City outbreak*. bioRxiv, 2021.

16. Bolger, A.M., M. Lohse, and B. Usadel, *Trimmomatic: a flexible trimmer for Illumina sequence data*. Bioinformatics, 2014. 30(15): p. 2114–20.

17. Li, H. and R. Durbin, *Fast and accurate short read alignment with Burrows-Wheeler transform*. Bioinformatics, 2009. 25(14): p. 1754–60.

18. Hadfield, J., et al., *Nextstrain: real-time tracking of pathogen evolution*. Bioinformatics, 2018. 34(23): p. 4121–4123.

19. Elbe, S. and G. Buckland-Merrett, *Data, disease and diplomacy: GISAID’s innovative contribution to global health*. Glob Chall, 2017. 1(1): p. 33–46.

20. Rambaut, A., et al., *A dynamic nomenclature proposal for SARS-CoV-2 lineages to assist genomic epidemiology*. Nat Microbiol, 2020. 5(11): p. 1403–1407.

21. Rambaut, A., et al., *Addendum: A dynamic nomenclature proposal for SARS-CoV-2 lineages to assist genomic epidemiology*. Nat Microbiol, 2021. 6(3): p. 415.

22. Zhao, Z., et al., *Moderate mutation rate in the SARS coronavirus genome and its implications*. BMC Evol Biol, 2004. 4: p. 21.

23. Worobey, M., et al., *The emergence of SARS-CoV-2 in Europe and North America*. Science, 2020. 370(6516): p. 564–570.

24. Cerutti, G., et al., *Potent SARS-CoV-2 neutralizing antibodies directed against spike N-terminal domain target a single supersite*. Cell Host Microbe, 2021. 29(5): p. 819–833 e7.

25. Gong, Y.N., et al., *SARS-CoV-2 genomic surveillance in Taiwan revealed novel ORF8-deletion mutant and clade possibly associated with infections in Middle East*. Emerg Microbes Infect, 2020. 9(1): p. 1457–1466.

26. Su, Y.C.F., et al., *Discovery and Genomic Characterization of a 382-Nucleotide Deletion in ORF7b and ORF8 during the Early Evolution of SARS-CoV-2*. mBio, 2020. 11(4).

27. Hachim, A., et al., *ORF8 and ORF3b antibodies are accurate serological markers of early and late SARS-CoV-2 infection*. Nat Immunol, 2020. 21(10): p. 1293–1301.

28. Pereira, F., *Evolutionary dynamics of the SARS-CoV-2 ORF8 accessory gene*. Infect Genet Evol, 2020. 85: p. 104525.

29. Young, B.E., et al., *Effects of a major deletion in the SARS-CoV-2 genome on the severity of infection and the inflammatory response: an observational cohort study*. Lancet, 2020. 396(10251): p. 603–611.

